# Distribution of informal caregiving for older adults living with or at risk of cognitive decline within and beyond family in rural South Africa

**DOI:** 10.1101/2024.06.20.24309077

**Authors:** Sostina S Matina, Lenore Manderson, Michelle Brear, Farirai Rusere, F. Xavier Gómez-Olivé, Kathleen Kahn, Guy Harling

**Affiliations:** MRC/Wits Rural Public Health and Health Transitions Research Unit, School of Public Health, University of the Witwatersrand, Johannesburg, South Africa; School of Public Health, University of the Witwatersrand, Johannesburg, South Africa; School of Social Sciences, Monash University, Melbourne, Australia; School of Public Health and Preventive Medicine, Monash University, Melbourne, Australia; School of Animal, Plant and Environmental Sciences, University of the Witwatersrand, Johannesburg, South Africa; Institute for Global Health, University College London, London, United Kingdom; Africa Health Research Institute, KwaZulu-Natal, South Africa; School of Nursing and Public Health University of KwaZulu-Natal, Durban, South Africa

**Author notes:** Address for correspondence: Guy Harling. Institute for Global Health, University College London, Mortimer Market Centre, London, WC1E 6JB. **Disclaimer**: The funders had no role in study design, data collection and analysis, decision to publish, or preparation of the manuscript. **Authors’ contributions**: LM and GH conceptualised the study, SM and GH conducted the analyses, and SM wrote the first draft of the paper. All authors contributed to the study design, data interpretation, and final revisions to the text.

**Keywords:** Caregiving, aging, familial care, informal care, South Africa

## Abstract

**Objectives:** Aging populations in rural areas of low and middle-income countries will increasingly need care. However, formal support is severely limited and adult children are frequently unavailable due to morbidity, early mortality, employment and migration. We aimed to describe how care is shared within and between households for older adults in a rural South African setting.

**Methods:** We conducted quantitative interviews with 1012 household members and non-household caregivers of 106 older adults living with or at risk of cognitive decline in rural Mpumalanga, South Africa. Using descriptive statistics and regression analysis, we described how care is shared, with particular attention to generational patterns of care.

**Results:** Informal care for older persons was spread among family, friends, and neighbours, with minimal paid support. This care was mostly provided by female relatives one or two generations younger than the recipient, and unemployed. However, a smaller number of paid caregivers, also mostly female, provided the most intensive care. Spouses commonly took on the role of primary caregiver for their partner.

**Discussion:** In our study, care mainly came from household members due to close family ties and practical considerations, with support from outside the household. This reflects shared history, reciprocal relationships, and easy access to care tasks within the household. A deeper understanding of how informal care for older adults is shared in low- and middle-income countries is essential for developing targeted interventions.

## BACKGROUND

With increased life expectancy in African countries, people are increasingly reporting chronic conditions significantly associated with older age (Guerchet et al., 2017; Olayinka & Mbuyi, 2014). In South Africa, older people experience multiple overlapping health conditions. These include living with long-term HIV, infectious and non-communicable diseases (NCDs), notably hypertension, diabetes, cancer, and vision and hearing impairments (Gómez-Olivé et al., 2017; Oni et al., 2014). The prevalence of cognitive decline and dementia is also rising in low- and middle-income countries (LMICs) as populations age (Hazzan et al., 2016; Mehta & Yeo, 2017). While all older adults will require care at times, caregiving for a person with dementia is particularly demanding because cognitive decline reduces individuals’ ability to live independently and interact socially (Adedeji et al., 2022; Cao et al., 2020)

Most care for people with dementia is provided informally at home, typically by unpaid relatives and friends of the care recipient. Dementia caregiving is commonly characterised as a dyadic process, with a single close family member providing near-continuous support. In practice, however, in both higher income and low-income settings, care is shared among multiple individuals (Koehly et al., 2015; Spillman et al., 2019), reflecting the complexity of care needs involved and the differing capacities of caregivers. In some high-income countries and settings this informal care is buttressed by formal support (e.g., home visits by nurses) (Hajek et al., 2016), but in LMICs available evidence indicates that dementia-related caregiving largely falls on family members (Kane et al., 2021). This reflects both cultural norms which dictate who should give care and generate perceptions that children who send their parents to care homes are selfish (Agbawodikeizu et al., 2023; Rutagumirwa et al., 2020), and very limited institutional settings or alternatives to informal care. For example, the South African public healthcare system uses an Integrated Chronic Disease Management (ICDM) model which focuses on health promotion, self-management, population screening, facility reorganisation, and clinical management support to meet population needs, but does not include dementia-specific services (Godongwana et al., 2021). As a result, those living with dementia, especially in rural areas, rely entirely on informal care, predominantly from female family members (Jacobs et al., 2023; Roomaney et al., 2023). This unequal caregiving is likely to exacerbate existing inequalities in chronic illness and multimorbidity amongst older South African women (National Department of Health (NDoH) & Council (SAMRC), 2017; Payne et al., 2017), given that caregivers of people living with dementia report more significant subjective burden and lower quality of life even than other caregivers (Karg et al., 2018).

Everyday care is generally provided in the care recipient’s home, in which case the primary caregiver may have moved residence, or in the caregiver’s home, in which case the care recipient may have been moved. However, LMIC households are frequently multigenerational – one-third of all South African households contained members at least two generations apart in 2011 (Makiwane, 2017) – and strongly connected to nearby extended family and friends (Kelly et al., 2017). In such circumstances, despite a key person usually bearing responsibility for care, caregiving may be shared among multiple people within a social network to limit the workload for any one person (Mortazavi et al., 2015). A larger caregiving network reduces morbidity and mortality, and increases medication adherence for care recipients (Knight & Schatz, 2022; Naslund et al., 2020). A wider range of caregivers also benefits care recipients who can thus receive both psychological social support to foster a sense of belonging and mitigate loneliness, and instrumental support for functional limitations, physical care, and medication adherence and healthcare service access.

The complex nature of those households complicates the sharing of care within and beyond households in southern Africa. This complexity includes missing generations due to e.g. HIV mortality (Roos & Wheeler, 2016; Schatz, 2018), and non-resident generations due to circular migration for employment, both of which reduce the availability of working age adults in the locations where older people receive care (Adamek et al., 2020; Nzima & Maharaj, 2020). These processes result in “skipped generation” households, in which the only resident members are young dependents and older people, usually grandparents and grandchildren or great-grandchildren. Ideas of reciprocity and intergenerational obligation dictate that grandchildren care for their aging grandparents (Nzima & Maharaj, 2020; Schatz, 2018). However, many of these grandchildren (and great-grandchildren) are adolescents whose capacity to care may be limited by educational commitments, age and other factors (Kidman & Thurman, 2014; Stols et al., 2016). Evidence on how these factors lead to the spread of caregiving within and beyond households is very limited.

We therefore evaluated how informal care provision for over 100 older adults with care needs in rural South Africa was distributed across their social networks, with particular attention to generational and kinship patterns and numbers of caregivers.

## METHODS

### Setting

Our study was nested within the Agincourt Health and Socio-Demographic Surveillance System (HDSS) site of the South African Medical Research Council/University of the Witwatersrand Rural Public Health and Health Transitions Research Unit, in Bushbuckridge, Mpumalanga Province, South Africa. The Agincourt HDSS has maintained an annual census of ∼120,000 people in 31 rural Mpumalanga communities since 1992 (Kahn et al., 2012). The area has limited employment opportunities in commercial food production and tourist venues (proximate to Kruger National Park), driving circular labour migration to urban areas. The area also experienced significant numbers of deaths from HIV in the 1990s and 2000s. These two factors jointly result in relatively small cohorts of working age adults and many skipped-generation households – around 11% of households have skipped generations within Agincourt HDSS, of which two-thirds include adults aged over 60 (Schatz et al., 2015). Public healthcare services are limited to three district hospitals 25 to 60kms away, to which many residents cannot afford to travel, and eight local primary healthcare facilities that lack specialist services.

### Study design

#### Participant selection and sampling strategy

The Agincourt HDSS provides a robust platform, including an established community and rigorous sampling frame for numerous epidemiological studies, including HAALSI (Health and Aging in Africa: A Longitudinal Study of an INDEPTH Community in South Africa), which commenced in 2014, enrolling a random sample of 5059 adults aged ≥40 (Gómez-Olivé et al., 2018). The HAALSI Dementia Study began in 2019–20 to estimate the prevalence and incidence of dementia and mild cognitive impairment (MCI) in the HAALSI cohort and conducted a second interview round in 2021 (see Bassil et al., 2022). The study received ethics approval from Wits Health Research Ethics Committee (Ethics number) and relevant South African government committees (withheld for review).

We sampled index cases (older people with or at risk of cognitive impairment, hereafter referred to as care recipients) from Wave 2 of the HAALSI Dementia Study (see Manderson et al., 2022). We first included all participants clinically diagnosed with moderate or severe dementia in Wave 1 and remained in the cohort at Wave 2. Since clinical diagnoses for Wave 2 were not available at the time of sampling, we identified additional potential index cases algorithmically, modelled on past work using similar methods to ascertain dementia status when clinical ratings are not available (Crimmins et al., 2011; Gianattasio et al., 2020).

First, we constructed a multinomial logit model using selected cognitive and informant variables from Wave 1 to predict a three-level dementia severity score based on consensus diagnoses at Wave 1: none; mild; moderate to severe. This model was 92% accurate in predicting Wave 1 clinical dementia diagnosis severity category. We then applied coefficients from the prediction model to the cognitive and informant measures collected in Wave 2, to predict probable Wave 2 clinical severity. By sampling design, care recipients were approximately evenly split between men and women. We sampled individuals from those with the highest predicted dementia severity until we had reached a total of 117 index cases; the final sample included 51 males and 55 females. Ethnographic data collected in 21 of these households during Kaya suggested that while those sampled did not all have an observable cognitive impairment, almost all care recipients received care daily.

For each care recipient, we first interviewed the designated primary household respondent from HAALSI Dementia Study Wave 2. They were asked to list all resident and non-resident household members plus any non-household kin or non-kin who provided care to the care recipient. We then conducted quantitative interviews with all household members and caregivers who were not household members, aged ≥12, who provided verbal informed consent (with prior consent from parents or guardians for all minors). We included minors aged 12–17 since they are likely to play an important role in caregiving, and having a household member living with cognitive impairment is likely to have a substantial impact on their wellbeing. Interviews were conducted face-to-face between July and December 2022 at respondents’ homes, with interviewers using tablet computers to capture data.

#### Measures

We investigated care distribution from both the caregivers’ and care recipients’ point of view. Our primary outcome at the caregiver level was self-reported average weekly hours of care provided. We also calculated the percentage of all care to a given recipient provided by each caregiver, based on average weekly hours of care. Caregivers were also asked if they considered themselves the primary caregiver to the care recipient and how confident they were in their capacity to provide care using a Likert scale (not at all confident; a little confident; somewhat confident; mostly confident; and extremely confident).

At the care recipient level, we calculated total weekly hours of care received by summing across all caregivers as well as the maximum duration in years that the recipient had been receiving care from any current caregiver. Concentration of care was measured using the Herfindahl-Hirschman index (HHI), which assesses how a variable is distributed among distinct entities (Brezina et al., 2016; Rój, 2018). HHI is calculated as Σs2, where s2 represents the share of the total amount each entity contributes and thus ranges from 0 to 1, with higher values indicating a greater concentration. We used reported weekly hours of caregiving from caregivers to calculate a care recipient-level measure of care concentration. We considered care recipient’s gender, age in decades (<70, 70-80, 80-89, ≥90) and their predicted cognitive impairment severity (no dementia; mild dementia; moderate-severe dementia). We also considered caregiver’s age (<18, 19-39, 40-59, ≥60), gender, marital status (married/coresident; never married; previously married), education (none; any primary, any secondary; any tertiary) and work status (fulltime work; part-time work; seeking work; out of workforce).

At the caregiver-care recipient dyad level, we considered the age difference between the care recipient and each caregiver in years, gender homophily (yes or no), relationship and household membership status. We categorised the relationship of caregiver to recipient at eight levels, five reflecting kinship and three others. Kin were distinguished as: spouse; non-spouse peer generation (sibling, cousin, in-law, etc) or older generations; child; non-child child generation (e.g., niece, nephew, daughter-in-law); grandchild and beyond (e.g., great niece, great-grandchild). Non-kin were separated into: employee/paid helper; friend; and neighbour. We also distinguished between those who were or were not members of the same household as the care recipient.

#### Statistical analyses

We first generated descriptive statistics using median and interquartile range (IQR) for continuous variables and percentages for categorical variables. We assessed differences by gender in exposures and outcomes using Pearson’s x2 tests for categorical variables and Wilcoxon rank-sum tests for continuous variables. We assessed care provision as experienced by the care recipient (i.e., total care per household) divided into eight relationship categories, and separately divided into three broader categories (resident relative; non-resident relative; non-relative), using proportions of all care and 95% confidence intervals (CI). We further subdivided these analyses by gender of responder. We calculated the age difference between potential caregivers and care recipients and presented this in histograms and violin plots, and calculated the gender homophily of care recipients and caregivers.

To assess the predictors of caregiver provision level, we used negative binomial regression analysis with outcomes of: i) caregivers’ hours of care; and ii) caregivers’ proportion of all care to their care recipient – the latter to remove confounding of associations by the overall quantity of care provided to each recipient. We conducted logistic regression with an outcome of caregivers reporting themselves to be the care recipient’s primary caregiver. Last, we ran an ordered logit regression with an outcome of reported confidence in providing care. All models were run as random intercept multilevel regressions (respondents nested in households), with covariates as listed in the exposures section. We used R version 4.3.2 for our statistical analyses (R Development Core Team, 2010).

## RESULTS

Care recipients resided in 24 villages. Of the 117 care recipients whose caregivers were sampled, two had passed away, one was unknown at the location, three were used to pilot the survey and one missing. The primary respondent in four households declined to participate. Among the remaining 106 care recipients to whom a primary respondent consented and was interviewed, 1020 household members and non-household members who provided care to the recipients were named, and 1012 of these subsequently consented and completed a survey (three outmigrated, one died, three were non-contactable and one declined). Over half of the care recipients were over 80 years of age (Table 1). Three-quarters of female care recipients (76%) were widowed, while nearly half of male recipients (49%) were married. Reflecting the larger numbers of women predicted to have poor cognition in the HAALSI Dementia Study; female care recipients were more likely to be predicted to have dementia. Two-thirds (67 of 106; 63%) of care recipients were reported to receive no formal care.ive

**Table 1:** Care recipient characteristics.

|  | **Overall** | **Male** | **Female** | **p-value<sup>1</sup>** |
| --- | --- | --- | --- | --- |
| N | 106 | 51 | 55 |  |
| Marital status |  |  |  | <0.001 |
| Married/coresident | 29.5% | 49.0% | 11.1% |  |
| Never married | 4.8% | 3.9% | 5.6% |  |
| Divorced | 8.6% | 9.8% | 7.4% |  |
| Widowed | 57.1% | 37.3% | 75.9% |  |
| Predicted Dementia |  |  |  | <0.001 |
| No dementia | 15.1% | 31.4% | 0.0% |  |
| Mild dementia | 67.0% | 58.8% | 74.5% |  |
| Moderate-severe dementia | 17.9% | 9.8% | 25.5% |  |
| Age group |  |  |  | 0.80 |
| Under 70 | 14.2% | 15.7% | 12.7% |  |
| 70-79 | 24.5% | 27.5% | 21.8% |  |
| 80-89 | 43.4% | 41.2% | 45.5% |  |
| 90+ | 17.9% | 15.7% | 20.0% |  |

All values are percent unless otherwise noted. <sup>1</sup> Pearson's Chi-squared test for categorical variables; Wilcoxon rank sum test for continuous variables; <sup>2</sup> Scale from 0-10 where values over 5 indicate worsening health.

In total, 978 of the 1012 respondents (96,8%) reported providing care, including 251 of 351 household members (71.5%). Respondents’ marital status varied, reflecting the wide range of life stages represented in the sample, with men more likely to be never or currently married and women more likely to be previously married (separated, divorced or widowed). Fewer than 30% of caregivers were working. Despite more than 60% of respondents being of working age, almost half of respondents were out of work and not looking for a job. Thirty-two of the 106 care recipients received care from their spouses. Figures for gender homophily suggested no preferential involvement in care based on the gender of the recipient.

The average care recipient received 52 hours of care weekly (IQR: 24, 70). The concentration of care varied widely, with a median HHI of 0.22 (IQR: 0.18, 0.31) across the 106 households (Figure 1). Only a handful of households had an HHI over 0.5, i.e., one individual provided more than 70% of all care hours. Caregivers provided care for a median of five years (IQR: 3, 10). The median of the longest dyad relationships per household was 13 years (IQR: 7, 20), and the longest dyadic care relationship was 40 years.

**Figure 1:**
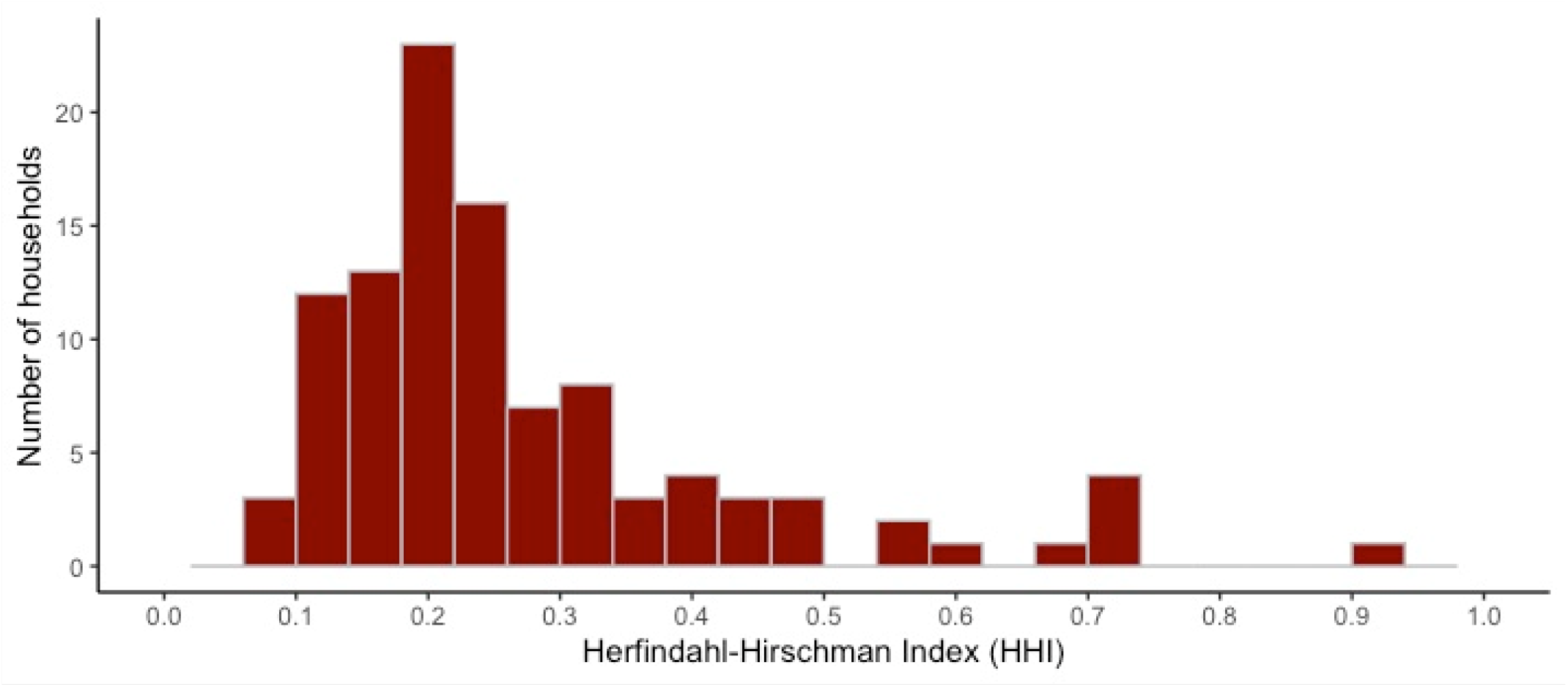
Concentration of care provision to care recipients

Care was shared among kin and non-kin caregivers (Supplementary Figure 4). The most common carers were children (25% of all caregivers) who provided a mean of 5 hours of care per week (95% confidence interval [CI]: 4, 6) each, or 14 hours (95%CI: 11, 17) in total (Figure 2 and Supplementary Table 1). Grandchildren provided a similar amount of care, comprising 23% of all caregivers and providing a mean of 6 hours of care a week each (95%CI: 5, 7), or 17 hours (95%CI: 12, 21) per household. However, when present in the caregiving network, non-kin paid and spousal caregivers provided more hours of care per week than children (mean 22; 95%CI: 14-30) and grandchildren – (mean 18;95%CI: 14-23). Female spouses and daughters provided a significantly greater proportion of care than their male counterparts even allowing for the greater number of female caregivers – i.e., women each provided more hours.

**Figure 2:**
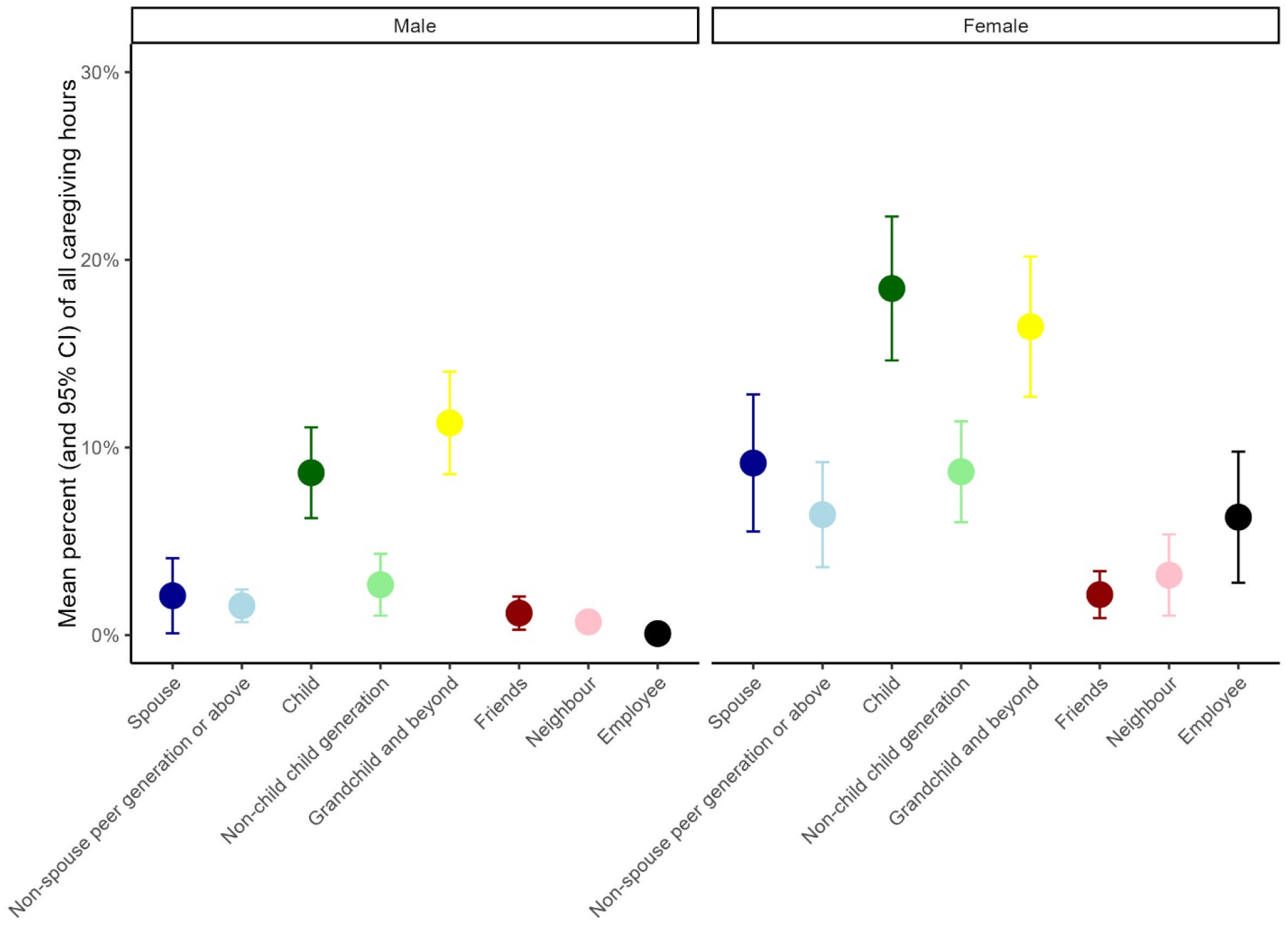
Distribution of caregivers hours from care recipient’s point of view based on relationship type and caregiver gender

Almost two-thirds of reported caregivers were female (Table 2). Most respondents were aged under 40, (59% of males vs 48% of females) leading to a slightly larger age gap between care recipients and their male caregivers (45 vs 41 years). Only 13.8% were under 18. The median age difference between caregivers and care recipients was 42 years (IQR: 29, 57), with almost 20% of caregivers over 40 years younger and very few older (Supplementary Figure 2). When stratified by relationship type (Figure 3), beside expected generational kin differences, almost all non-kin paid caregivers were female and around 40 years younger than their care recipients. Friends were typically closer in age than neighbours to the care recipient. When caregivers were stratified by relative and residence status, resident kin provided the greatest proportion of all care, followed by non-resident kin and then non-kin; females in all groups reported providing more hours of care each week than males (Supplementary Figure 3).

**Figure 3:**
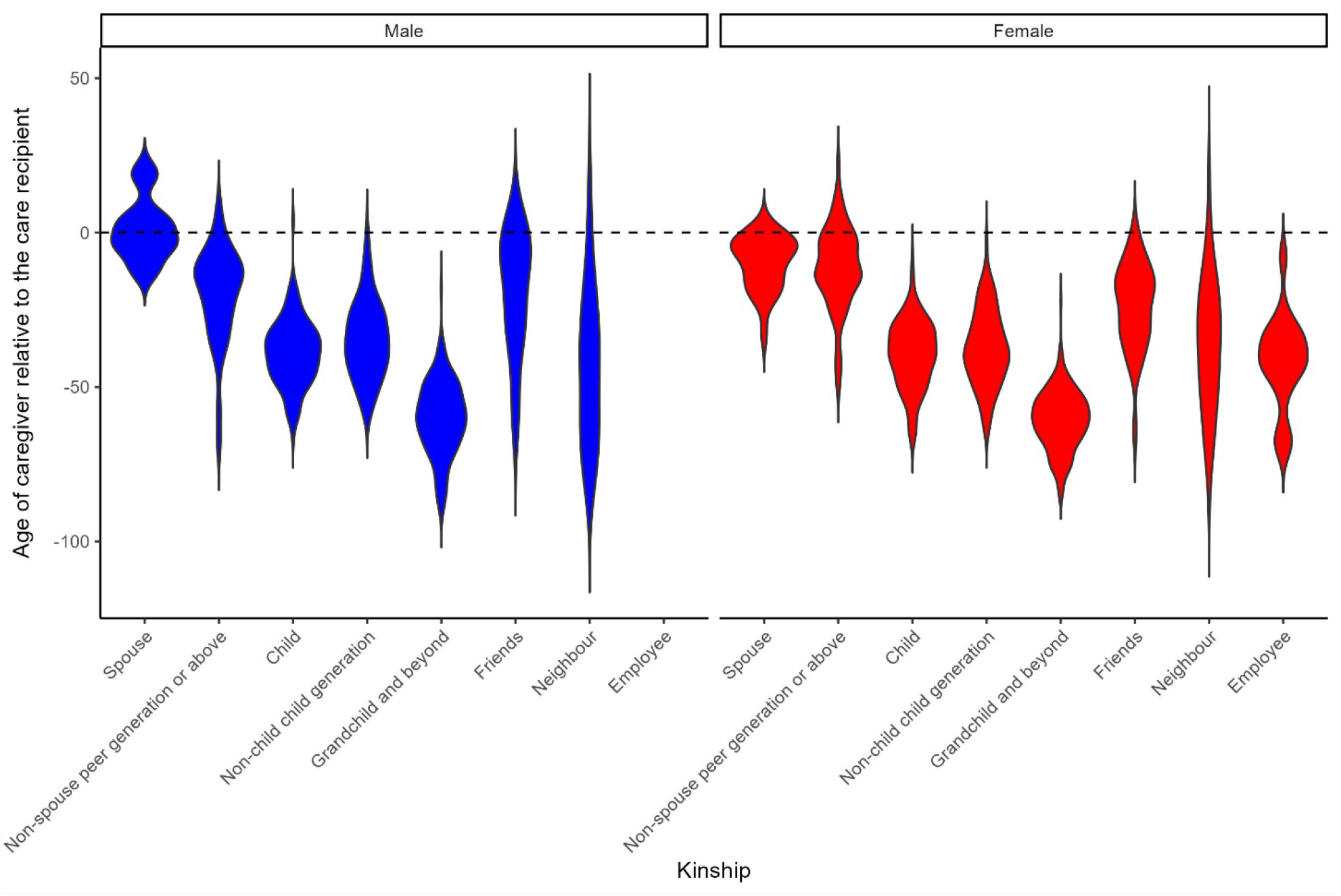
Age difference between caregiver and care recipient by relationship type and caregiver gender

**Table 2:** Respondents characteristics.

|  | Overall | Male | Female | p-value<sup>1</sup> |
| --- | --- | --- | --- | --- |
| N | 1012 | 398 | 614 |  |
| Caregiver Age, % |  |  |  | 0.007 |
| Under 18 | 13.8% | 14.1% | 13.7% |  |
| 19-39 | 38.0% | 43.7% | 34.4% |  |
| 40-59 | 30.6% | 27.6% | 32.6% |  |
| 60+ | 17.5% | 14.6% | 19.4% |  |
| Marital status, % |  |  |  | <0.001 |
| Married/coresident | 36.5% | 38.9% | 34.9% |  |
| Never married | 42.5% | 48.2% | 38.8% |  |
| Previously married | 21.0% | 12.8% | 26.4% |  |
| Number of living children, Median (IQR) | 2 (0, 4) | 2 (0, 3) | 2 (1, 4) | 0.001 |
| Work status, % |  |  |  | <0.001 |
| Fulltime work | 19.8% | 27.6% | 14.7% |  |
| Part-time work | 9.6% | 11.8% | 8.1% |  |
| Seeking work | 22.5% | 19.1% | 24.8% |  |
| Out of workforce | 48.1% | 41.5% | 52.4% |  |
| Age difference, Median (IQR) | 42 (29, 56) | 45 (32, 58) | 41 (27, 55) | 0.003 |
| Gender homophily, % |  |  |  | <0.001 |
| Both female | 30.2% |  | 49.8% |  |
| Male recipient, female provider | 30.4% |  | 50.2% |  |
| Female recipient, Male provider | 20.5% | 52.0% |  |  |
| Both male | 18.9% | 48.0% |  |  |
| Relation status, % |  |  |  | <0.001 |
| Spouse | 3.3% | 1.5% | 4.4% |  |
| Kin non-spouse peer generation or above | 9.3% | 8.5% | 9.8% |  |
| Child | 28.0% | 32.2% | 26.4% |  |
| Kin non-child child generation | 13.5% | 10.3% | 15.6% |  |
| Grandchild and beyond kin | 30.7% | 36.2% | 27.2% |  |
| Friends | 4.5% | 3.8% | 5.0% |  |
| Neighbour | 7.7% | 6.5% | 8.5% |  |
| Employee | 1.9% | 0.3% | 2.9% |  |
| Resident status, % |  |  |  | 0.008 |
| Relation - resident | 34.3% | 38.4% | 31.6% |  |
| Relation - non-resident | 51.4% | 51.0% | 51.7% |  |
| Non-relative | 14.2% | 10.6% | 16.6% |  |
| Education, % |  |  |  | <0.001 |
| None | 13.5% | 8.5% | 16.8% |  |
| Primary Education | 19.8% | 20.1% | 19.5% |  |
| Secondary Education | 56.5% | 58.5% | 55.2% |  |
| Tertiary Education | 10.2% | 12.8% | 8.5% |  |

All values are percent unless otherwise noted. <sup>1</sup>Pearson's Chi-squared test for categorical variables; Wilcoxon rank sum test for continuous variables

### Predictors of caregiving

In multivariable models, household members provided significantly more hours of care (incidence rate ratio [IRR]: 2.17, 95%CI: 1.58-2.75) and a higher proportion of care (IRR: 1.77, CI: 1.45-2.09) than non-members (Table 3). Respondents aged over 60 provided the most hours of care, and those aged 19-39 were the fewest, but there was little difference by age in percentage of care provided, implying that older respondents were in households with more overall care provision. Degree of employment was negatively associated with care provision – those seeking work provided around twice as much care as those in full-time work. Levels of care provision were higher both from female respondents and to female care recipients – the highest care levels were seen when both individuals were female. Each decade of age difference between caregiver and recipient was associated with an approximately 10% decrease in both hours and percentage of care provided. As descriptive analysis shows, employees and spouses provided the most intensive care, while neighbours, same-generation kin and friends provided the least.

**Table 3:** Multilevel regression models to predict Caregiver Provision Level.

|  | Hours of care | Percentage of care |
| --- | --- | --- |
| Household member vs non-member | 2.17 [1.58, 2.75] | 1.77 [1.45, 2.09] |
| Caregiver age (vs ≥ 60) |  |  |
| < 18 | 0.88 [0.15, 1.61] | 1.03 [0.46, 1.60] |
| 19-39 | 0.60 [0.20, 1.01] | 1.07 [0.57, 1.56] |
| 40-59 | 0.72 [0.42, 1.01] | 1.12 [0.68, 1.55] |
| Marital status (vs Married) |  |  |
| Never married | 1.06 [0.77, 1.35] | 1.03 [0.80, 1.27] |
| Previously married | 0.88 [0.68, 1.07] | 0.97 [0.71, 1.22] |
| Work status (vs full-time work) |  |  |
| Part-time work | 1.00 [0.68, 1.32] | 1.45 [1.01, 1.90] |
| Seeking work | 2.00 [1.41, 2.59] | 2.05 [1.44, 2.66] |
| Out of workforce | 0.89 [0.60, 1.18] | 1.38 [0.98, 1.79] |
| Gender homophily (vs Both male) |  |  |
| Male recipient, female provider | 1.24 [0.94, 1.54] | 1.20 [0.98, 1.42] |
| Female recipient, Male provider | 1.18 [0.74, 1.63] | 0.94 [0.71, 1.16] |
| Both female | 1.88 [1.16, 2.60] | 1.55 [1.16, 1.94] |
| Age difference (per decade) | 0.94 [0.80, 1.08] | 0.90 [0.82, 0.98] |
| Relationship type (vs Spouse) |  |  |
| Non-spouse peer generation or above | 0.43 [0.22, 0.64] | 0.33 [0.17, 0.49] |
| Child | 0.66 [0.32, 1.00] | 0.44 [0.22, 0.67] |
| Non-child child generation | 0.64 [0.30, 0.99] | 0.44 [0.22, 0.67] |
| Grandchild and beyond | 0.81 [0.27, 1.35] | 0.53 [0.25, 0.81] |
| Friends | 0.52 [0.21, 0.83] | 0.35 [0.18, 0.52] |
| Neighbour | 0.36 [0.11, 0.60] | 0.25 [0.12, 0.38] |
| Employee | 2.76 [0.84, 4.68] | 1.86 [0.82, 2.90] |
| Education (vs Primary) |  |  |
| No Education | 1.47 [0.87, 2.07] | 0.93 [0.64, 1.22] |
| Secondary Education | 0.84 [0.65, 1.03] | 0.81 [0.62, 0.99] |
| Tertiary Education | 0.84 [0.51, 1.17] | 0.55 [0.34, 0.75] |
| Care recipient predicted dementia (vs no dementia) |  |  |
| Mild dementia | 0.63 [0.33, 0.93] | 0.85 [0.67, 1.03] |
| Moderate to severe dementia | 0.86 [0.42, 1.31] | 0.96 [0.67, 1.26] |

Values are incidence rate ratios and 95% confidence intervals.

Household members were substantially more likely to report that they were primary caregivers compared to non-members (Odds ratio [OR]: 3.69; 95%CI: 2.28-5.96), as were those closer in age to recipients, females, and those with tertiary education (Table 4). These patterns were largely repeated for those who were primary caregivers by hours provided, although the associations with household membership and female gender were stronger, as was the association between fulltime employment and less caregiving primacy. Caregiver confidence was higher for those with tertiary education (OR: 2.69; 95%CI: 1.45-4.98), those who had been caregivers for longer and those who provided more hours of care per week.

**Table 4:**
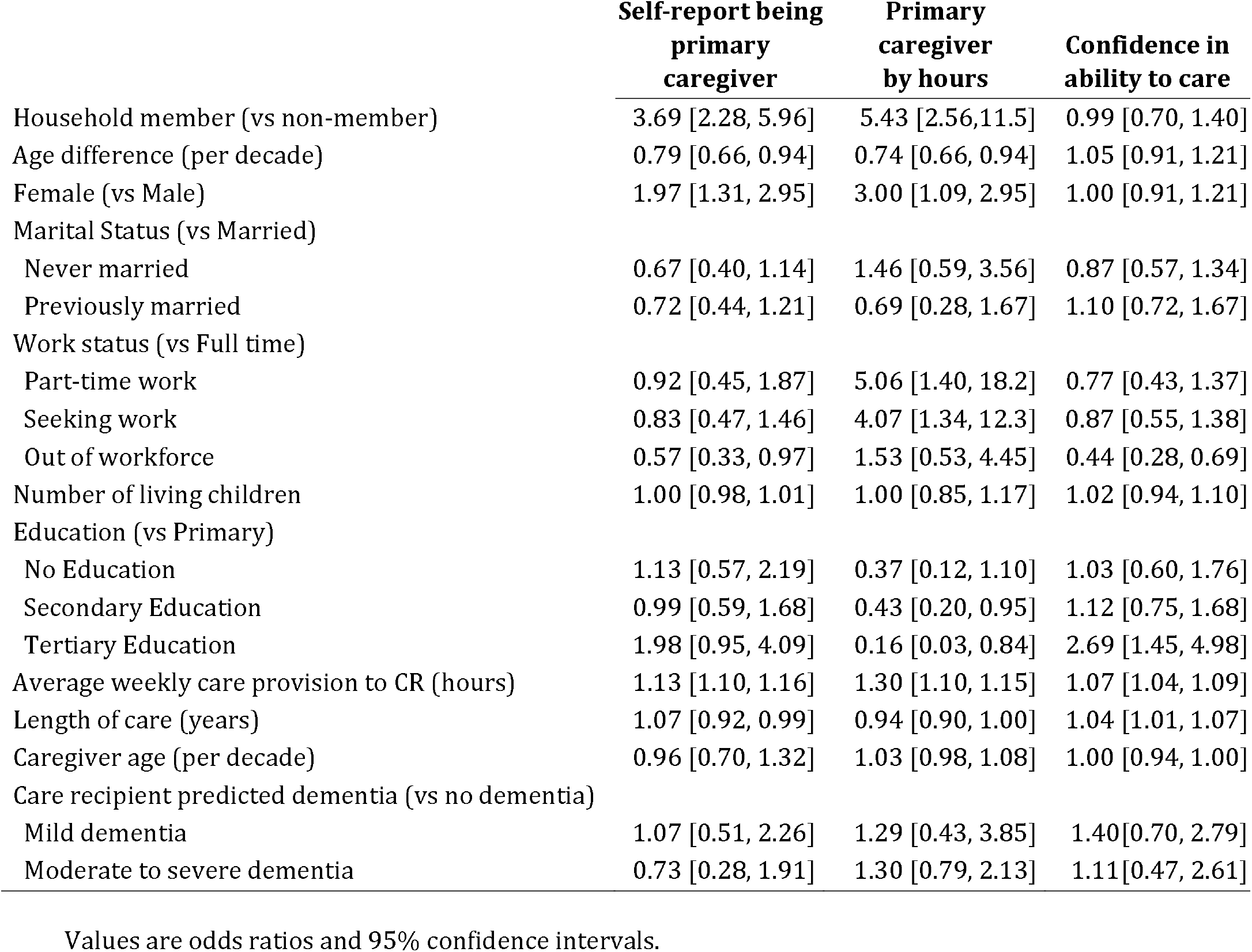
Multivariable regression models to predict caregiving outcomes.

## DISCUSSION

In this rural South African setting, informal care for older persons with care needs was based widely on family, friends and neighbours – with very limited paid caregiving and almost no formal care. Care was most commonly provided by female relatives one or two generations younger than the recipient and unemployed. However, a smaller number of spouses and paid caregivers provided the most intensive care. One-third of our care recipients had spousal carers, with female spouses frequently reporting being primary caregivers; male spouses were rarer, reflecting large marital age gaps and higher female life expectancy (and thus higher mortality) and probably also gender norms. This spousal primacy is common globally (Bainbridge et al., 2006; Park et al., 2015), and may reflect either a strong marital commitment, cultural expectation to care for one’s spouse in times of need (Bohman et al., 2009), or both. In some cases, inheritance, shared assets (e.g house), and continued access to support from families may also have influenced the decision to continue to provide care. While gender-imbalanced spousal care may reflect available support capacity, greater numbers of female carers and more intensive care from females were consistent across relationship types. These patterns reflect gender norms that expect women to assume primary caregiving roles; such caregiving, in turn, may perpetuate gender inequalities in labour force participation and other opportunities (Roos & Wheeler, 2016; Rutagumirwa et al., 2020). Furthermore, gender-inequitable care puts particular pressure on female-headed households, which are common in South Africa in part due to high male labour migration, AIDS-related and NCDs mortality (Nwosu & Ndinda, 2018; Nzima & Maharaj, 2020). This dual role as caregiver and provider can further exacerbate women’s financial struggles and increase their vulnerability to poverty.

While children commonly care for older adults worldwide, reflecting intertemporal reciprocity of care that may reflect self-interested or altruistic motivations (Silverstein et al., 2002), we found grandchildren and great-grandchildren contributing significantly to caregiving in rural Mpumalanga. There are at least three possible, likely overlapping, explanations for the frequency of this ‘skipped generation’ care. First, it may reflect direct reciprocity in a setting where many grandparents raised their grandchildren due to HIV-related mortality and economic migration (Adamek et al., 2020; Schatz et al., 2018). Second, as noted above, grandchild care roles may also reflect the financial support older relatives provide to them via the Older Person’s Grant a means-tested non-contributory monthly pension paid to people aged over 60 years which is often a primary income source in poor South African households (Waidler & Devereux, 2019). Third, grandchild care may reflect opportunity and convenience as youth, unable to find work, are living and waiting for opportunities in rural homes (Honwana, 2014). Centrally, the multigenerational households prevalent in rural South Africa offer a built-in support system for caregiving, allowing care responsibilities to be shared among family members (Ardington et al., 2010). This is especially important for ensuring older people receive care in the context of limited resources and social support systems, but also places financial strain on older people and their households (Schatz et al., 2018).

Household members provided more care than non-household members in our sample. This pattern reflects both within-family propinquity – i.e., shared history, reciprocity, mutual understanding and emotional bonds – and pragmatics – small acts of care are easily anticipated and undertaken when caregiver and care recipient are together, and thus care tasks expand where the caregiver and care recipient are proximate (Adedeji et al., 2022; Schatz et al., 2018). Nevertheless, care recipients received substantial support from beyond the household, including care from an average of five non-resident family members and around 1.5 non-family members. The presence of geographically and socially close-knit communities in rural villages provides access to wider support networks, further influencing caregiving practices and resource-sharing, such as driving care recipients to the clinics (Schatz et al., 2018). Nineteen care recipients had paid caregivers, but these employees provided the most intensive care of any group. The limited availability of paid care might reflect a desire from relatives to give care themselves – potentially linked to ideas of reciprocity and social exchange within families (Akinrolie et al., 2020; Schatz, 2018). However, it also reflects the very limited resources available within care recipients’ households, many of which depend almost exclusively on government grants – only 19% had fulltime employment.

### Strengths and limitations

Our findings provide an overview of the distribution of caregiving for older people in one rural area of South Africa. The nesting of our study within an existing cohort (HAALSI)(Bassil et al., 2022) with over a decade of data providing rich background information on the care recipients, and our response rate was very high. By casting a wide net, our study captured many actual and potential caregivers of older individuals, providing a comprehensive view of their care support network. However, the study also has limitations. Its cross-sectional design means we did not capture the dynamic nature of caregiving over time. Longitudinal studies are needed to examine how caregiving dynamics evolve and impact caregivers’ and care recipients’ well-being, something the linked qualitative data may be able to inform in the future. Our use of self-reported caregiving quantity and primacy may have been affected by systematic over-reporting (males were more likely to report being primary caregivers despite providing fewer hours of care on average than women).

## Conclusion

The distribution of care given to older people in rural South Africa is complex. Care is typically shared amongst a range of household residents, non-residents kin and non-kin. As in high income country settings, caregiving is gendered; women provide more care than men. However, in our setting, younger people who were grandchildren or great-grandchildren more often cared for older people than children. While we have shown how care is shared, the motivations behind this pattern of care, and their dynamics over time, require additional investigation. Such deeper understanding is essential for developing targeted interventions and support systems to address the caregiving needs of vulnerable populations – both care recipients and providers – to promote greater gender equity in caregiving responsibilities.

## Data Availability

All data produced in the present study are available upon reasonable request to the authors

## Acknowledgements

We thank the research assistants who conducted the interviews and the study respondents for their participation. The research on which this article is based is nested within the MRC/Wits Rural Public Health and Health Transitions Research Unit and Agincourt Health and Socio-Demographic Surveillance System, a node of the South African Population Research Infrastructure Network (SAPRIN), supported by the Department of Science and Innovation, the University of the Witwatersrand, the Medical Research Council, South Africa, and previously The Wellcome Trust (058893/Z/99/A; 069683/Z/02/Z; 085477/Z/08/Z; 085477/B/08/Z). This research was funded in whole, or in part, by the Wellcome Trust [Grant number 210479/Z/18/Z]. For the purpose of open access, the author has applied a CC BY public copyright licence to any Author Accepted Manuscript version arising from this submission.

## SUPPLEMENTARY MATERIALS

**Supplementary Table 1:** Mean and 95% confidence interval of care hours provided by relationship status.

|  | Individual hours | Total hours |
| --- | --- | --- |
| Spouse | 18 [14, 23] | 5 [3, 8] |
| Non-spouse peer generation or above | 3 [3,6] | 4 [2, 5] |
| Children | 5 [4, 6] | 14 [11, 17] |
| Non-child child generation | 2 [0,6] | 5 [4, 7] |
| Grandchild and beyond | 6 [5, 7] | 17 [12, 21] |
| Friends | 4 [3, 6] | 2 [1, 3] |
| Neighbour | 3 [2,4] | 2 [1, 4] |
| Employee | 22 [14, 30] | 4 [2, 6] |

Individual hours refers to mean hours per caregiver in the group; Total hours refers to all hours by all caregivers within this group.

**Supplementary Figure 1:**
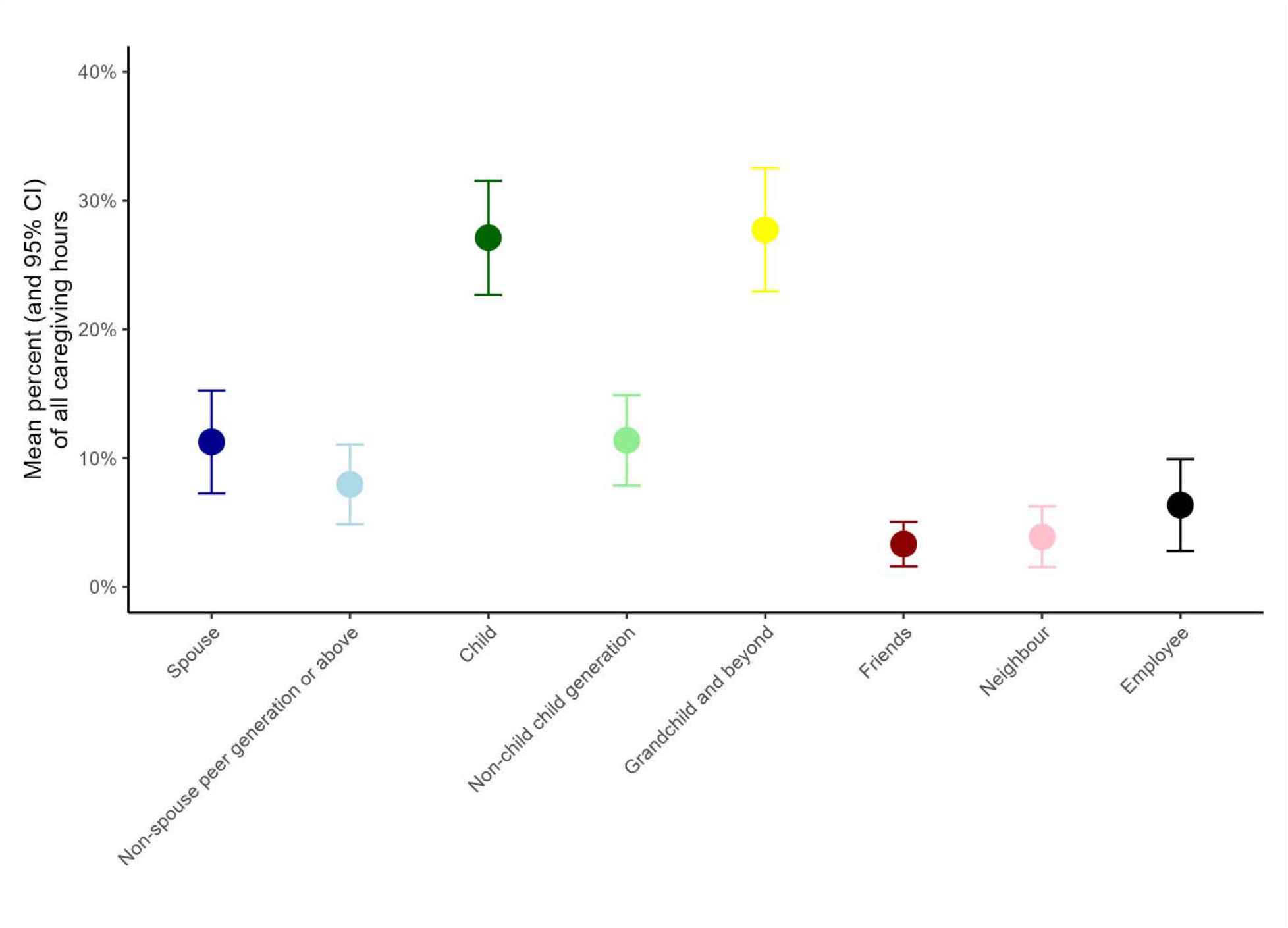
Distribution of caregiver hours from the care recipient’s point of view based on relationship type

**Supplementary Figure 2:**
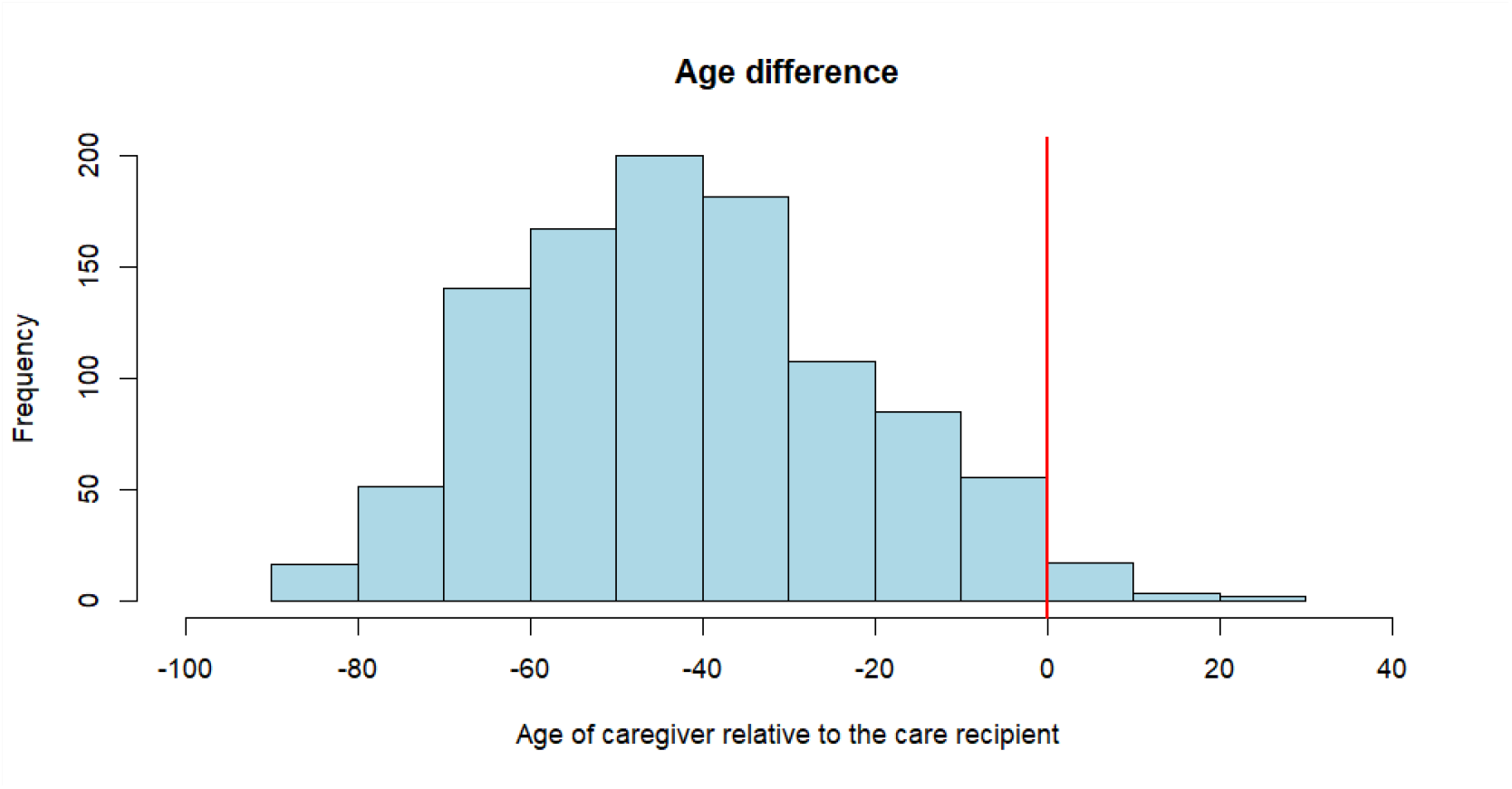
Age difference between care recipient and caregiver (negative values mean caregiver is younger)

**Supplementary Figure 3:**
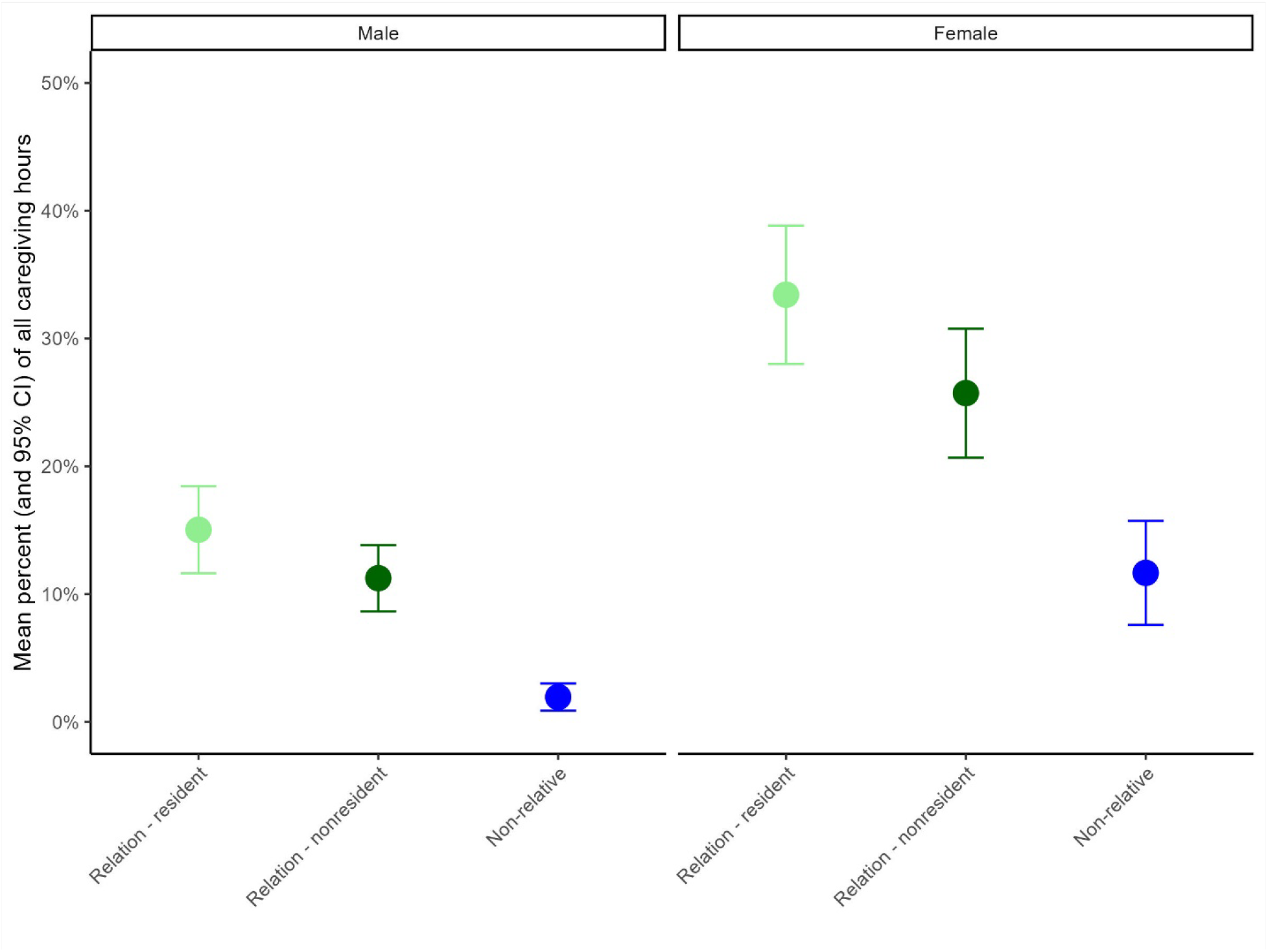
Distribution of caregiver hours from care recipient’s point of view based on household membership status and gender

**Supplementary Figure 4:**
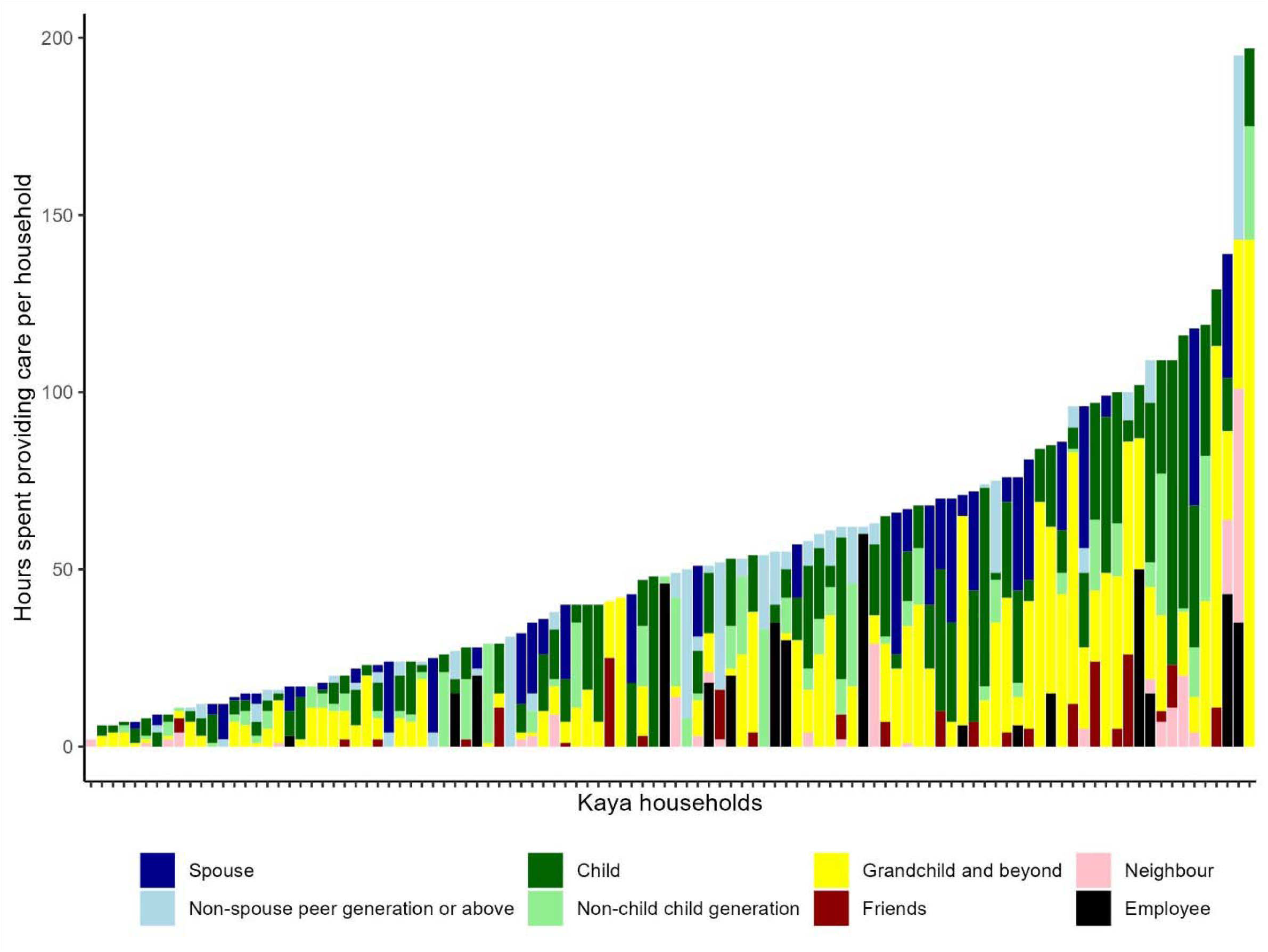
Composition of hours of care provided for each care recipient by relationship type

